# Independent validation of the PrecivityAD2™ blood test to identify presence or absence of brain amyloid pathology in individuals with cognitive impairment

**DOI:** 10.1101/2025.04.05.25325203

**Authors:** Justine Coppinger, Tim West, Kris M. Kirmess, Ilana Fogelman, Sutapa Ray, Sheena Aurora, Philip B. Verghese, Joel B. Braunstein, Kevin E. Yarasheski, the Alzheimer’s Disease Neuroimaging Initiative (ADNI)

## Abstract

**Objective:** The diagnostic performance of the Amyloid Probability Score 2 (APS2) - the algorithmic result of the PrecivityAD2™ blood test - was originally trained and validated in two cohorts of cognitively impaired (CI) individuals. Using an independent cohort to evaluate blood test reliability, we conducted an external diagnostic accuracy assessment of the validated APS2 cut point as it is currently applied in clinical practice.

**Methods:** Plasma biomarker ratios Aβ42/40 and p-tau217/np-tau217 (expressed as %p-tau217) were quantified and incorporated into the APS2 algorithm in samples obtained from 192 Alzheimer’s Disease Neuroimaging Initiative participants with CI (70% mild cognitive impairment / 30% dementia). APS2 diagnostic performance was determined using amyloid positron emission tomography (PET) as the reference standard. Plasma biomarkers were quantified in a CLIA-certified, CAP-accredited laboratory (C2N Diagnostics, St. Louis, MO) using liquid chromatography-tandem mass spectrometry.

**Results:** APS2 values were significantly higher in the 56% of CI participants with a positive amyloid PET scan. Concordance with amyloid PET was high (AUC-ROC 0.95 (95%CI): 0.93– 0.98); 54% of participants had a positive APS2. The previously validated APS2 cut point yielded an overall accuracy of 91% (95%CI: 86-94%), sensitivity 90% (95%CI: 83-94%) and specificity 92% (95%CI: 84-96%).

**Interpretation:** The PrecivityAD2 blood test’s APS2 identified brain amyloid pathology with accuracy, sensitivity, and specificity ≥ 90% in this intended use population. This external validation reaffirms the diagnostic robustness of this blood biomarker test and supports its use as a confirmatory test, consistent with published expert recommendations, for assessment of presence or absence of brain amyloid pathology in symptomatic patients.

## Introduction

Globally, Alzheimer’s disease (AD) affects an estimated 25-30 million people and is a leading cause of disability and death[1–3]. Identifying the presence of brain amyloid plaques and neurofibrillary tau tangles early in the pathophysiological processes that characterize AD is crucial for establishing an accurate and early diagnosis as well as determining timely eligibility for therapeutic interventions that slow disease progression and cognitive decline.

Novel blood biomarker (BBM) tests for diagnosis of AD pathology are currently being developed, validated, and compared[4–22]. Recently, AD BBM key opinion leaders and expert panels convened by the World Health Organization (WHO), Global CEO Initiative on AD (CEOi), and the Alzheimer’s Association, outlined diagnostic performance characteristics that should be considered with fluid biomarkers targeted for use in clinical settings[8,12,23–26]. Blood biomarker tests that demonstrate high performance compared to conventional brain amyloid diagnostic tools such as amyloid positron emission tomography (PET) or cerebrospinal fluid (CSF) biomarkers can complement cognitive testing, rapidly aid in ruling in or ruling out an AD diagnosis in symptomatic patients, and accelerate the diagnostic process for more timely initiation of a disease-modifying treatment[12,25]. Further, given the complexities of biomarker variability and the implications of diagnostic thresholds, repeated validation studies and reliability testing are necessary for evaluating robustness, accuracy, and generalizability.

C2N Diagnostics’ PrecivityAD2™ blood test is a multianalyte assay with algorithmic analysis that uses immunoprecipitation, liquid chromatography-tandem mass spectrometry (LC-MS/MS) to quantify plasma concentrations of amyloid beta 42 (Aβ42) and Aβ40 isoforms, and phosphorylated-tau217 (p-tau217) and non-phosphorylated-tau217 (np-tau217) peptides. Plasma Aβ42/40 and p-tau217/np-tau217 (%p-tau217) concentration ratios are calculated and incorporated into an algorithm that generates the Amyloid Probability Score 2 (APS2) - a value that ranges from 0 to 100 and represents the likelihood that brain amyloid pathology is present or absent in individuals with CI[9,15,27]. Prior studies have analytically and clinically validated PrecivityAD2 analytes and APS2 cut points against amyloid PET imaging and CSF biomarkers of brain amyloid pathology[15,16,28–32].

In a recent head-to-head round-robin comparison of BBM tests for AD pathology in cognitively impaired and unimpaired participants, a study developed and launched by the Foundation for the NIH (FNIH) Biomarkers Consortium, plasma samples from the Alzheimer’s Disease Neuroimaging Initiative (ADNI) biorepository were analyzed using the leading amyloid and p-tau tests, each employing different analytical platforms. A prespecified analytic plan was developed for a head-to-head comparison of these plasma biomarkers against amyloid PET as the primary outcome[16]. The FNIH study analyzed the performance of the C2N analytes individually and when combined (%p-tau217, p-tau217, Aβ42/40) into a *de novo* prediction model, and found that the model outperformed the other BBM models for identifying brain amyloid pathology status. However, the diagnostic accuracy of the APS2 algorithm in the intended use population was not determined in that study for two reasons: the algorithm validation was not a prespecified study objective; and the ADNI cohort included cognitively unimpaired participants, which falls outside of the intended use population for the PrecivityAD2 test.

The purpose of the present study was to externally validate the prespecified APS2 algorithm and cut point used in clinical care in the FNIH-ADNI cohort of individuals with CI (i.e., the intended use population) (**Fig. 1**).

**Figure 1.**
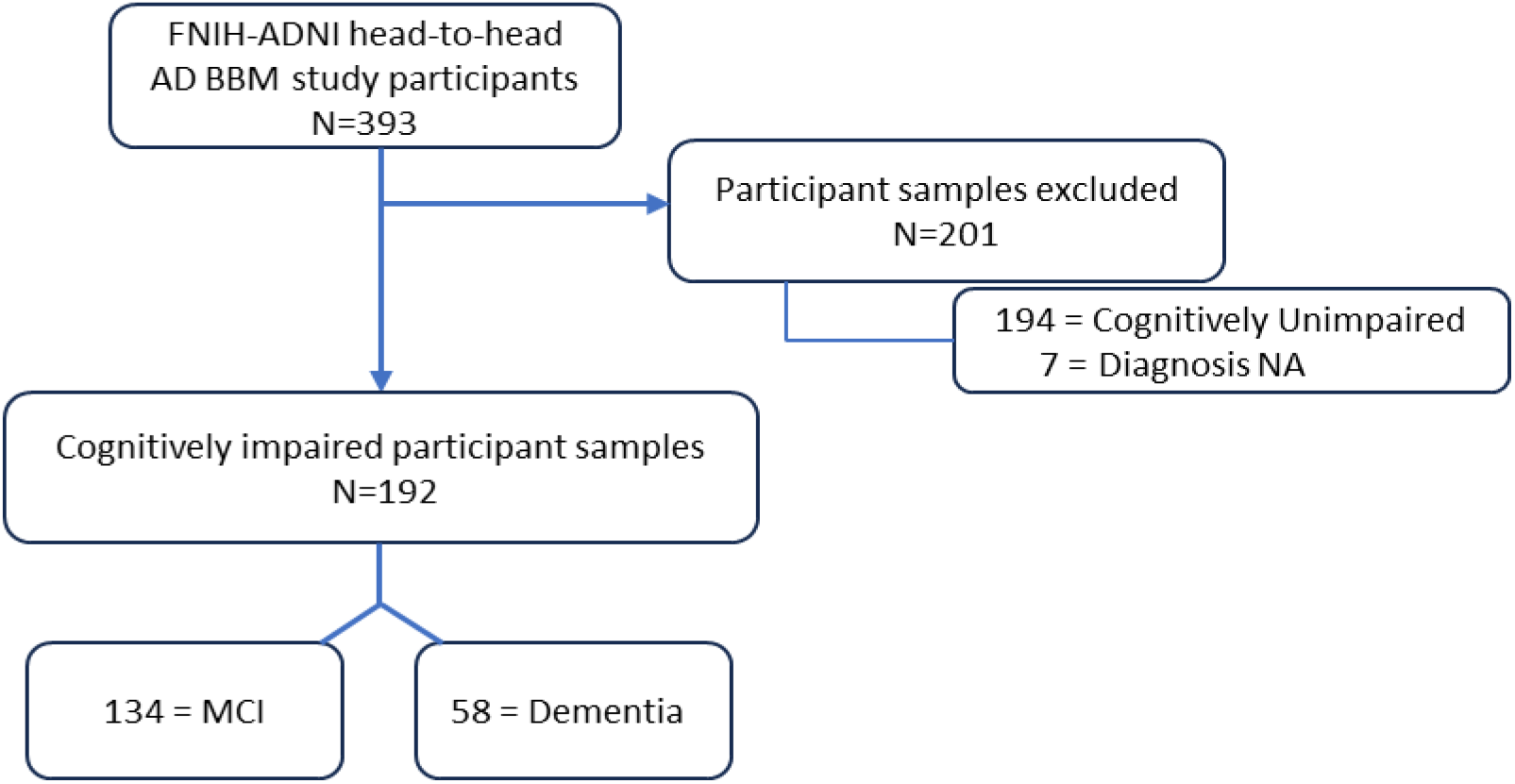
CONSORT diagram showing participant selection. From a total of 393 FNIH-ADNI participant samples[16], 201 with diagnosis assigned as “cognitively unimpaired” or “without diagnosis” data, were excluded. The remaining 192 participants with an “MCI” or “Dementia” diagnosis were included in the current analysis.

## Methods and Participants

### Participants

All ADNI volunteers received an explanation of study purpose, risks, and benefits, and all participants or their legally authorized representatives signed an IRB-approved consent document for their deidentified study related data to be shared with qualified researchers.

### ADNI Cohort selection

The FNIH project evaluated several candidate plasma biomarkers and compared their ability to identify amyloid PET status in cognitively impaired and unimpaired participants. The current analysis focused on the PrecivityAD2 test’s ability to identify amyloid PET status in 192 CI ADNI participants - 134 with mild cognitive impairment (MCI) and 58 with dementia - who had K_2_ EDTA plasma samples collected within six months of their most recent amyloid PET scan (**Fig 1**). All data analyzed herein, including demographic information (self-reported race, ethnicity, education, CDR, and sex), was downloaded from the ADNI database (https://adni.loni.usc.edu). ADNI was launched in 2003 as a public-private partnership, led by Principal Investigator Michael W. Weiner, MD. The original goal of ADNI was to test whether serial magnetic resonance imaging (MRI), PET, other biological markers, and clinical and neuropsychological assessments can be combined to measure the progression of MCI and early AD. The current goals include validating biomarkers for clinical trials, improving the generalizability of ADNI data by increasing diversity in the participant cohort, and providing data concerning the diagnosis and progression of AD to the scientific community.

### Amyloid PET status assignment

Standardized protocols for ^18^Florbetapir or ^18^Florbetaben amyloid PET imaging were used at ADNI sites. FreeSurfer v7.1 was used to identify the frontal, cingulate, parietal, and lateral temporal cortices, and to quantify the global standardized uptake value ratio (SUVR). SUVR was normalized to the whole cerebellum. The ADNI PET Core applied standardized data transformations to the ^18^Florbetapir and ^18^Florbetaben images and estimated the standardized global cortical Aβ burden in Centiloid (CL) units. For the current analysis, the presence of brain amyloid PET pathology was defined as CL > 25.

### PrecivityAD2 methods and APS2 algorithm

The analytical methods for PrecivityAD2 biomarkers and the clinical validation of the APS2 algorithm have been described previously[15,28–32]. Briefly, blood was collected into K_2_ EDTA vacutainers^®^, plasma was isolated and stored according to ADNI protocols. Frozen plasma aliquots were shipped on dry ice from the ADNI biorepository to the C2N Diagnostics laboratory (St. Louis, MO USA) for processing and LC-MS/MS analysis of the PrecivityAD2 biomarkers. These biomarkers were incorporated into the prespecified APS2 algorithm. All C2N lab personnel involved in sample analysis remained blinded to all participant meta-data.

### Statistical analysis

All data analysis was performed using R version 4.3.1 (The R Foundation for Statistical Computing). Participant characteristics were summarized across the groups using means, medians, and standard deviations for continuous variables and using counts and percentages for categorical variables. Fisher’s exact test was used to compare accuracy observed for the different biomarkers. Receiver operating characteristic (ROC) analyses, including the calculation of the area under the ROC curve (AUC), sensitivity, specificity, positive predictive value (PPV), and negative predictive value (NPV) were conducted using the pROC package for R[33]. The 95% confidence intervals on accuracy, sensitivity, specificity, PPV and NPV were calculated using Wilson’s method[34].

## Results

This study included ADNI participants representing the intended use population for the PrecivityAD2 test: men and women above 55 years old with MCI or dementia (**Table 1**). The prevalence of amyloid PET positivity in this cohort was 56%, and 54% of the participants had a positive APS2. On average (**Supplementary Table 1 & Fig. 2A**), plasma Aβ42/40 was significantly lower in the amyloid PET positive group (0.0883 ± 0.0093) compared to the amyloid negative group (0.0983 ± 0.0107; *p* = 1.8e^−10^) and the area under the receiver operating characteristic curve (AUC-ROC) was 0.78 (95%CI: 0.71-0.85). Using the test’s prespecified plasma Aβ42/40 ratio cut point (0.095) gave an accuracy of 74% (95%CI: 68-80%), sensitivity 81% (95%CI: 73-88%), specificity 65% (95%CI: 55-75%), PPV 75% (95%CI: 66–82%), and NPV 73% (95%CI: 62–82%), values consistent with previous reports [30,31].

**Table 1.**
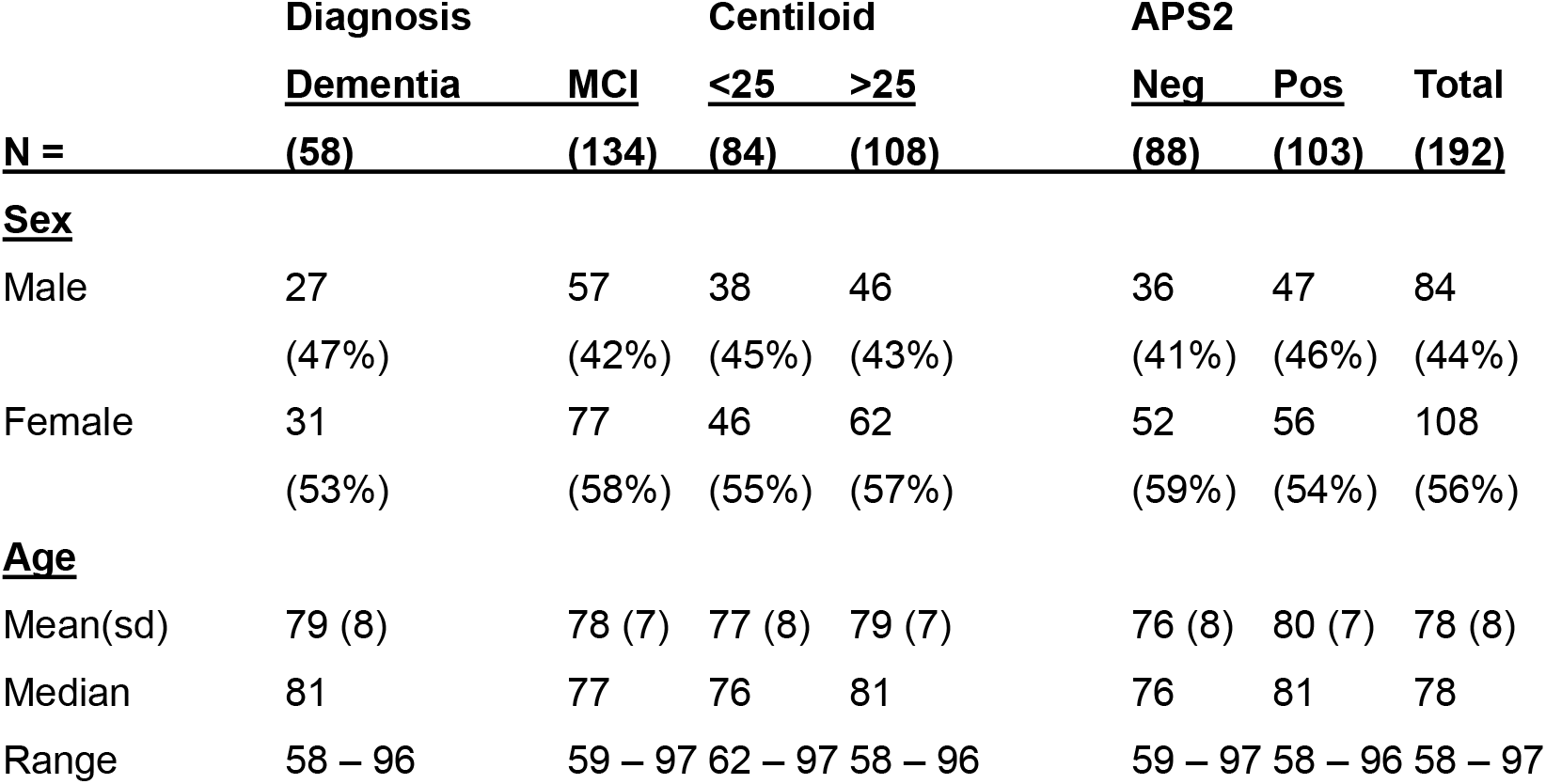
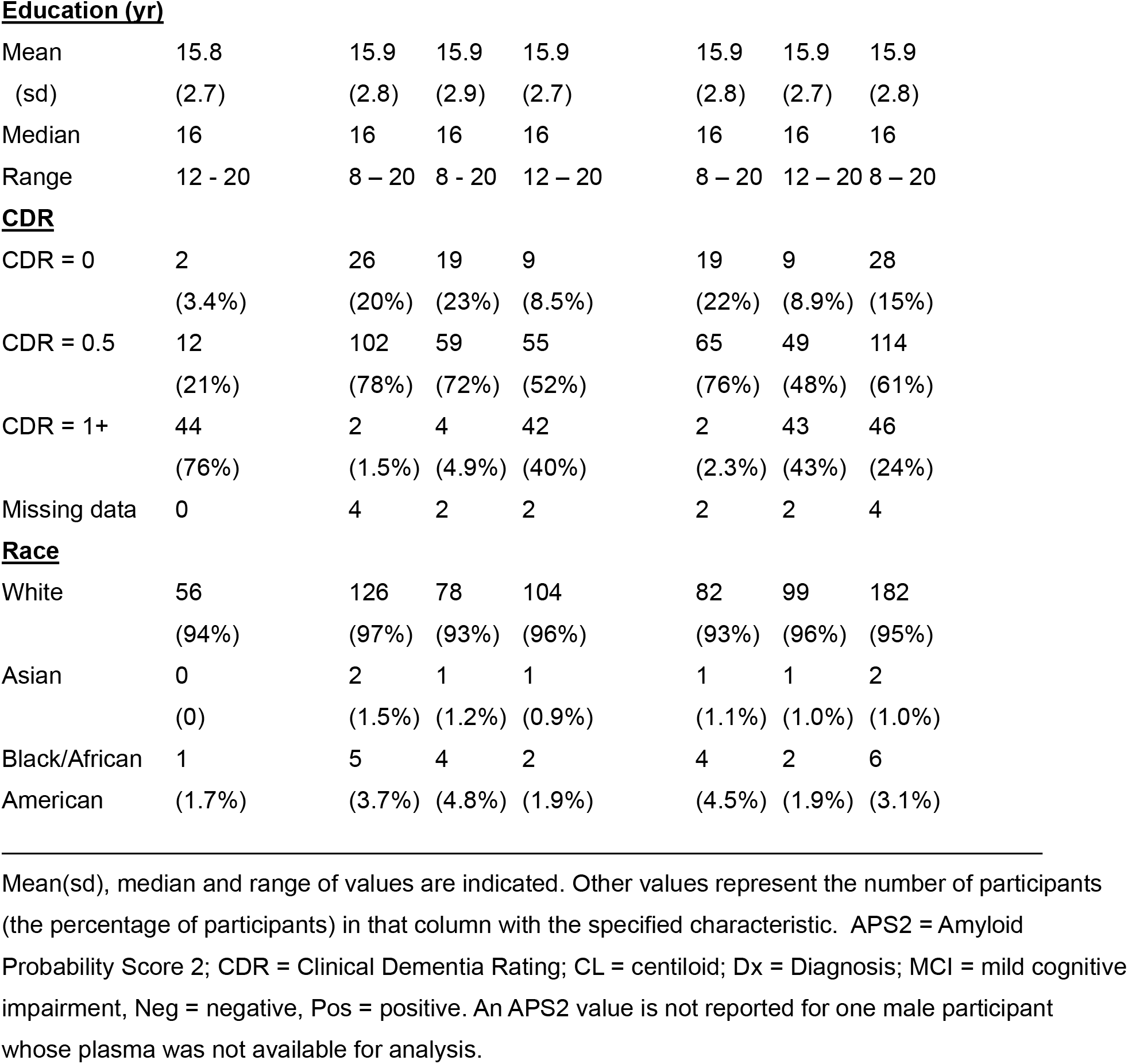
Demographics for 192 ADNI participants with either MCI or dementia.

|  | **Diagnosis** |  | **Centiloid** |  | **APS2** |  | **Total** |
| --- | --- | --- | --- | --- | --- | --- | --- |
|  | **Dementia** | **MCI** | **<25** | **>25** | **Neg** | **Pos** |  |
| **N =** | **(58)** | **(134)** | **(84)** | **(108)** | **(88)** | **(103)** | **(192)** |
| **<u>Sex</u>** |  |  |  |  |  |  |  |
| Male | 27<br>(47%) | 57<br>(42%) | 38<br>(45%) | 46<br>(43%) | 36<br>(41%) | 47<br>(46%) | 84<br>(44%) |
| Female | 31<br>(53%) | 77<br>(58%) | 46<br>(55%) | 62<br>(57%) | 52<br>(59%) | 56<br>(54%) | 108<br>(56%) |
| **<u>Age</u>** |  |  |  |  |  |  |  |
| Mean(sd) | 79 (8) | 78 (7) | 77 (8) | 79 (7) | 76 (8) | 80 (7) | 78 (8) |
| Median | 81 | 77 | 76 | 81 | 76 | 81 | 78 |
| Range | 58 – 96 | 59 – 97 | 62 – 97 | 58 – 96 | 59 – 97 | 58 – 96 | 58 – 97 |
| **Education (yr)** |  |  |  |  |  |  |  |
| Mean | 15.8 | 15.9 | 15.9 | 15.9 | 15.9 | 15.9 | 15.9 |
| (sd) | (2.7) | (2.8) | (2.9) | (2.7) | (2.8) | (2.7) | (2.8) |
| Median | 16 | 16 | 16 | 16 | 16 | 16 | 16 |
| Range | 12 - 20 | 8 – 20 | 8 - 20 | 12 – 20 | 8 – 20 | 12 – 20 | 8 – 20 |
| **CDR** |  |  |  |  |  |  |  |
| CDR = 0 | 2 | 26 | 19 | 9 | 19 | 9 | 28 |
|  | (3.4%) | (20%) | (23%) | (8.5%) | (22%) | (8.9%) | (15%) |
| CDR = 0.5 | 12 | 102 | 59 | 55 | 65 | 49 | 114 |
|  | (21%) | (78%) | (72%) | (52%) | (76%) | (48%) | (61%) |
| CDR = 1+ | 44 | 2 | 4 | 42 | 2 | 43 | 46 |
|  | (76%) | (1.5%) | (4.9%) | (40%) | (2.3%) | (43%) | (24%) |
| Missing data | 0 | 4 | 2 | 2 | 2 | 2 | 4 |
| **Race** |  |  |  |  |  |  |  |
| White | 56 | 126 | 78 | 104 | 82 | 99 | 182 |
|  | (94%) | (97%) | (93%) | (96%) | (93%) | (96%) | (95%) |
| Asian | 0 | 2 | 1 | 1 | 1 | 1 | 2 |
|  | (0) | (1.5%) | (1.2%) | (0.9%) | (1.1%) | (1.0%) | (1.0%) |
| Black/African | 1 | 5 | 4 | 2 | 4 | 2 | 6 |
| American | (1.7%) | (3.7%) | (4.8%) | (1.9%) | (4.5%) | (1.9%) | (3.1%) |

Mean(sd), median and range of values are indicated. Other values represent the number of participants (the percentage of participants) in that column with the specified characteristic. APS2 = Amyloid Probability Score 2; CDR = Clinical Dementia Rating; CL = centiloid; Dx = Diagnosis; MCI = mild cognitive impairment, Neg = negative, Pos = positive. An APS2 value is not reported for one male participant whose plasma was not available for analysis.

**Figure 2.**
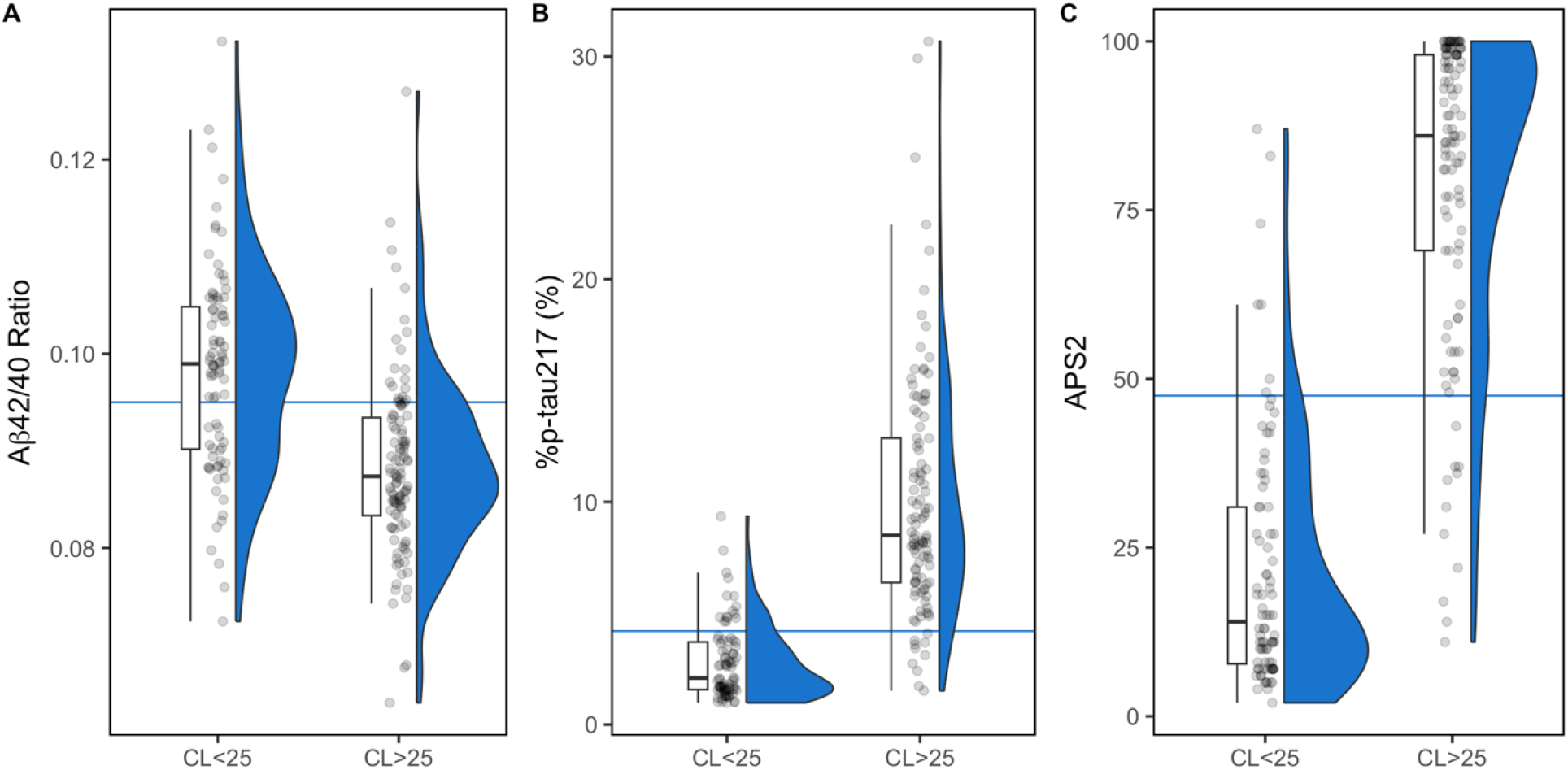
Combined box/whisker plots and distribution density plots for individual plasma **(A)** Aβ42/40, **(B)** %p-tau217, and **(C)** APS2 values. Boxes represent the upper and lower quartiles, the horizontal line in the boxes represents the median value, and the whiskers define the range of measured values. Plasma Aβ42/40 was significantly lower **(A)** and plasma %p-tau217 was significantly higher **(B)** in the CI participants with amyloid PET CL > 25 (i.e., presence of amyloid pathology). In **(A)**, the blue line represents the test’s prespecified Aβ42/40 cut point = 0.095. **(B)** The blue line represents the test’s prespecified %p-tau217 cut point = 4.2%. **(C)** APS2 values were higher for ADNI participants with amyloid PET CL > 25. The blue line represents the test’s prespecified APS2 cut point = 47.5.

On average, plasma %p-tau217 was significantly higher in the amyloid positive group (10.02 ± 5.40) than in the amyloid negative group with an AUC-ROC of 0.94 (95%CI: 0.92–0.98) (2.80 ± 1.69; *p* = 2.2e^−16^, **Supplementary Table 1 & Fig. 2B**). Using the test’s prespecified plasma %p-tau217 cut point (4.2%), yielded an accuracy of 87% (95%CI: 81-91%), sensitivity 91% (95%CI: 84-95%), specificity 82% (95%CI: 73-89%), PPV 87% (95%CI: 79-92%) and NPV 87% (95%CI: 78-93%), values consistent with previous reports [15,27].

The APS2 algorithm uses both the plasma Aβ42/40 and %p-tau217 values to calculate a likelihood score between 0 - 100. In the original CI validation cohort[15], an APS2 value of 47.5 was chosen as the cut point for distinguishing the presence of brain amyloid pathology from the absence. In the current study, APS2 was significantly higher in the amyloid PET positive group (83.8 ± 16.5) compared to the amyloid negative group (18.6 ± 12.8; *p =* 2.2e^−16^) and the AUC-ROC was 0.95 (95%CI: 0.93–0.98, **Supplementary Table 1 & Fig. 2C**). Using the test’s prespecified APS2 cut point, accuracy was 91% (95%CI: 86–94%), sensitivity 90% (95%CI: 83–94%), specificity 92% (95%CI: 84–96%), PPV 93% (95%CI: 87–97%) and NPV 88% (95%CI: 79–93%) (**Table 2**), values similar to those previously reported[15]. APS2 diagnostic accuracy at the prespecified cut point was better than that for plasma Aβ42/40 (74% vs. 91%, p < 0.0001), and numerically better than that for plasma %p-tau217 (87% vs. 91%, p = 0.33).

**Table 2.**
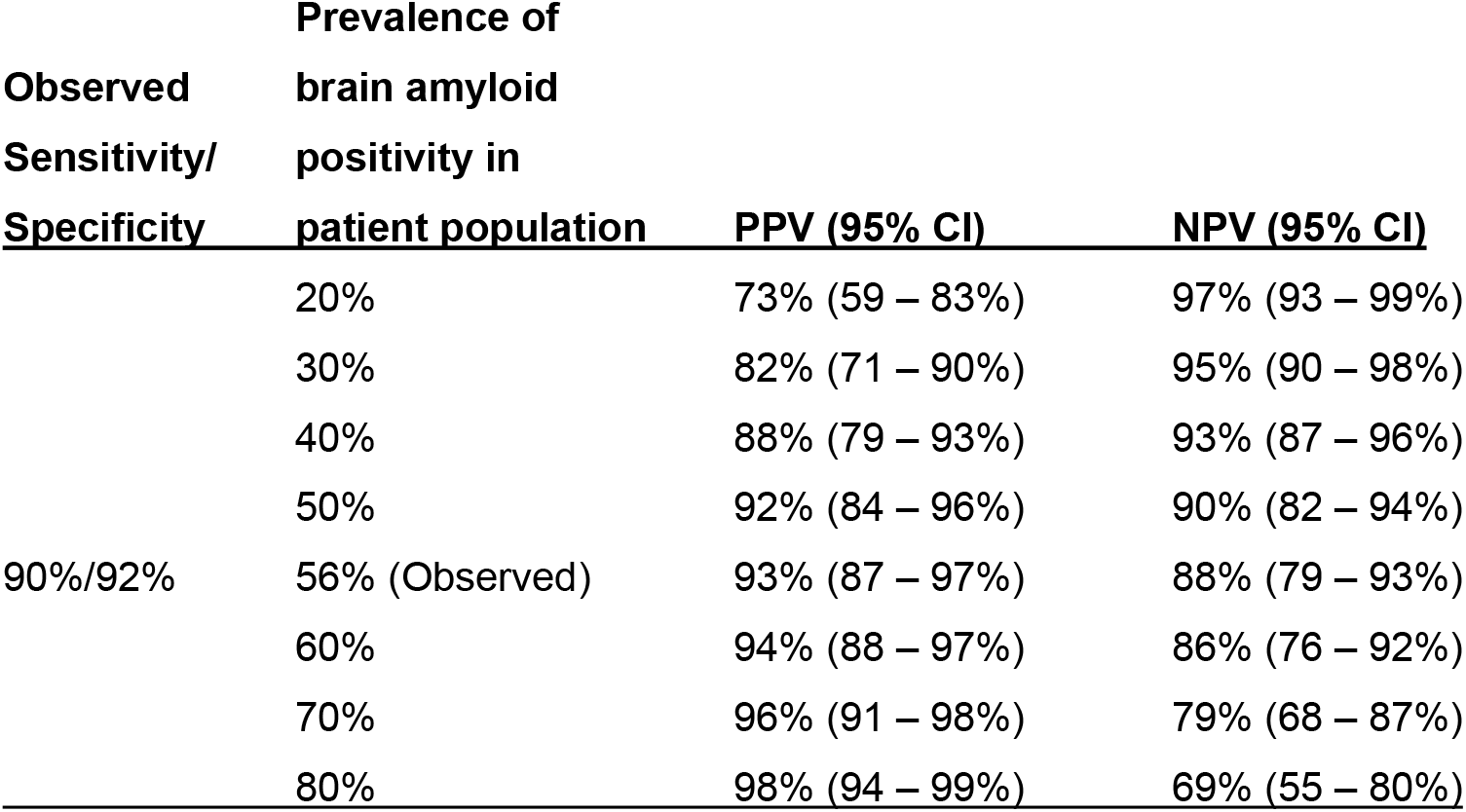
PrecivityAD2™ APS2 algorithm corresponding positive predictive and negative predictive values by prevalence of disease.

| **Observed**<br>**Sensitivity/**<br>**Specificity** | **Prevalence of**<br>**brain amyloid**<br>**positivity in**<br>**patient population** | **PPV (95% CI)** | **NPV (95% CI)** |
| --- | --- | --- | --- |
| 90%/92% | 20% | 73% (59 – 83%) | 97% (93 – 99%) |
|  | 30% | 82% (71 – 90%) | 95% (90 – 98%) |
|  | 40% | 88% (79 – 93%) | 93% (87 – 96%) |
|  | 50% | 92% (84 – 96%) | 90% (82 – 94%) |
|  | 56% (Observed) | 93% (87 – 97%) | 88% (79 – 93%) |
|  | 60% | 94% (88 – 97%) | 86% (76 – 92%) |
|  | 70% | 96% (91 – 98%) | 79% (68 – 87%) |
|  | 80% | 98% (94 – 99%) | 69% (55 – 80%) |

Based upon the observed sensitivity and specificity of the APS2 algorithm in this study, we also examined the impact of varying prevalence of amyloid pathology on the PrecivityAD2 test’s PPV and NPV (**Table 2**). This analysis is impactful in the context of individualized patient care where patient risk of disease influences confidence to rely on a diagnostic test to rule in or rule out disease. The observed prevalence of amyloid pathology in this study was 56%, consistent with an MCI / early dementia population, with an observed PPV and NPV of 93% and 88%, respectively. Among a predominantly asymptomatic, aged adult population (e.g., 20% - 30% prevalence of brain amyloid), the APS2 provides a very high NPV with point estimates of 95%-97%, highlighting its value for ruling out disease. Among a population with dementia (e.g., 70-80% prevalence of brain amyloid), the APS2 provides a very high PPV with point estimates of 96%-98%, highlighting its value for ruling in disease.

## Discussion

The findings in this independent cohort confirm the high diagnostic performance of the previously validated and prespecified single APS2 cut point to distinguish CI individuals with brain amyloid pathology from those without amyloid pathology. The current findings add clinically relevant information to the assay performance reported in the head-to-head study of Schindler and colleagues[16] given that APS2 is reported to and used by physicians as a diagnostic aid for their symptomatic patients. The observed PrecivityAD2 test accuracy, sensitivity, and specificity values from the current study, and calculated PPV and NPV percentages at different patient population prevalences are consistent with prior clinical validation data[15]. Further, they meet the current, recommended performance standards for confirmatory diagnostic BBM tests to identify amyloid pathology based on accuracy, sensitivity, and specificity values using a single cut point in the intended use population[12,24–26]. For an MCI / early dementia patient population with assumed 50% AD prevalence the APS2 algorithm and cut point showed ≥90% accuracy, with sensitivity and specificity values corresponding to 92% PPV and 90% NPV (**Table 2**).

Continued evaluations of test performance in independent populations, where patient heterogeneity and variation in sample collection methods can be substantial, enable healthcare providers to understand a test’s robustness, i.e., how the test will perform in their own patient population[35]. When clinical validity is assured, healthcare providers expect that a biomarker test for brain amyloid will have a direct impact on patient care by aiding in the diagnosis of the cause of cognitive decline and by guiding patient management[36].

Clinical utility studies have found that the PrecivityAD2 test improved physicians’ certainty in making clinically meaningful changes in AD/non-AD diagnoses, treatment appropriateness decisions including when to prescribe or not prescribe anti-AD therapies, and referral patterns for brain amyloid PET imaging[9,37]. PrecivityAD2 testing also helped physicians rule out AD, allowing for other diagnostic considerations[37–39]. A recent survey by the Alzheimer’s Association found that 70% of Americans want to know early if they have AD to allow for earlier interventions[40]. Early and accurate AD diagnosis can lead to positive patient outcomes, such as living independently for longer and maintaining a good quality of life because of access to appropriate services and support[41]. Other potential benefits may include more time for the patient and family to prepare, adjust and set realistic expectations for patient function, mood, and behaviors. For healthcare providers, early diagnosis may allow for more time to manage and treat underlying co-morbidities, address AD risk factors and safety issues, make referrals when necessary, and develop a comprehensive clinical management plan (e.g., existing or novel drugs, enroll in a clinical trial)[41].

Possible explanations for the reliability and replicability of the performance metrics of the PrecivityAD2 test’s APS2 algorithm include the use of a highly specific, high-resolution LC-MS/MS analytical platform for quantifying disease-specific peptides/proteins, and rigorous optimization and validation of plasma sample preparation methods in compliance with CLIA and CAP laboratory standards. In comparison to single-analyte immunoassays, this BBM test leverages the established analyte multiplexing capabilities and analyte specificity of LC-MS/MS to detect and quantify multiple Aβ or tau peptide species in each sample preparation and analytical workflow, which reduces inter-assay variability. This optimizes the quantitation of low abundant, plasma peptide species reflective of AD brain pathology. Variability is also reduced with the implementation of biomarker concentration ratios (rather than single analyte concentration alone) that can normalize for potential non-AD related factors expressed at the individual patient level. Variations in biomarker transport rates, blood volume, and biomarker metabolism have been shown to affect patient-specific AD biomarker levels, as have circadian rhythm, medications and differing lifestyle practices[42–44]. Presence of comorbidities, a common finding in individuals with AD, can also impact blood biomarker concentrations[45,46]. Higher plasma concentrations of Aβ42, Aβ40, p-tau217 and np-tau217 have been observed with chronic kidney disease, a condition that affects 40-50% of adults with AD[47,48]. Both Aβ42/40 and %p-tau217 are less impacted by chronic kidney disease compared to single-analyte biomarkers as concentration measures of both numerator and denominator for each set of biomarkers are thought to be similarly affected by alterations in renal clearance[48–51].

Although this replication cohort demonstrated diversity in patient age and sex (58-97 years, 50% women), the cohort is limited by the high education level (mean 16 years) and low number of under-represented minority individuals. These demographics highlight the cohort selection bias inherent to participants who have volunteered for enrollment into ADNI. Further, given the relatively small sample size, planned future evaluations will include larger, real-world cohorts that also include CI participants from varied social, educational and demographic backgrounds.

## Conclusion

The findings indicate that using the clinically validated PrecivityAD2 blood test’s APS2 algorithm in an independent population of CI individuals provided ≥90% accuracy, sensitivity, and specificity, consistent with prior clinical validation data. These findings provide further evidence of the replicability and reliability of the test’s APS2 algorithm and single cut point for identifying brain amyloid status in the intended use population. These results also support the test’s use as an alternative confirmatory diagnostic to CSF or amyloid PET biomarker assessments in accordance with established recommendations for clinical implementation.

## Data Availability

The datasets used and analyzed can be downloaded from the ADNI database (http://www.adni.loni.usc.edu) and the Parkinsons Progression Markers Initiative (PPMI) database (https://www.ppmi-info.org/access-data-specimens/download-data), RRID:SCR_006431.

http://www.adni.loni.usc.edu

## Author Contributions

All authors reviewed and edited initial manuscript drafts and reviewed and approved the final manuscript version. The study was conceptualized and executed by JC, TW, KMK, IF, SR, SA, PBV, JBB, and KEY. PBV supervised and KMK developed, monitored, and conducted quality control assessments during the plasma sample analysis at C2N Diagnostics. TW downloaded ADNI demographic and imaging data and provided independent statistical analysis.

## Availability of data and materials

The datasets used and analyzed can be downloaded from the ADNI database (http://www.adni.loni.usc.edu) and the Parkinson’s Progression Markers Initiative (PPMI) database (https://www.ppmi-info.org/access-data-specimens/download-data), RRID:SCR_006431.

## Acknowledgments

We thank all ADNI participants and their care partners. FNIH Biomarkers Consortium procured the ADNI plasma samples.

## Abbreviations

Aβ42: amyloid beta-42 (pg/mL)
Aβ40: amyloid beta-40 (pg/mL)
Aβ42/40: amyloid beta-42/amyloid beta-40 concentration ratio
AD: Alzheimer’s disease
ADNI: Alzheimer’s Disease Neuroimaging Initiative
APS2: Amyloid Probability Score 2
AUC: area under the curve
BBM: blood biomarker
CAP: College of American Pathologists
CDR: Clinical Dementia Rating
CEOi: Global CEO Initiative on Alzheimer’s disease
CI: cognitive impairment
CL: centiloid
CLIA: Clinical Laboratory Improvement Amendments
CSF: cerebrospinal fluid
K_2_ EDTA: dipotassium ethylenediamine tetraacetic acid
FNIH: Foundation for the National Institutes of Health
IVD: *in vitro* diagnostics
LC-MS/MS: liquid chromatography-tandem mass spectrometry
LDT: laboratory developed tests
MCI: mild cognitive impairment
MRI: magnetic resonance imaging
NA: no value available
NaN: value cannot be calculated
np-tau217: non-phosphorylated tau at threonine 217 (pg/mL)
NPV: negative predictive value
p-tau217: phosphorylated tau at threonine 217 (pg/mL)
%p-tau217: phosphorylated tau217/non-phosphorylated tau217 concentration ratio
PARIS: **P**lasma Test for **A**myloidosis **RI**sk **S**creening study
PET: positron emission tomography
PPV: positive predictive value
ROC: receiver operating characteristic curve sd standard deviation
SUVR: global standardized uptake value ratio
95%CI: 95% confidence interval
WHO: World Health Organization

