## Supplementary Table 1.1 for "Independent validation of the PrecivityAD2™ blood test to identify presence or absence of brain amyloid pathology in individuals with cognitive impairment"

| ***Supplementary Table 1.*** Plasma Aβ42/40, %p-tau217, and APS2 values for 192 ADNI participants with either MCI or dementia | | | | | | | | | |
| --- | --- | --- | --- | --- | --- | --- | --- | --- | --- |
|  | | **Dementia Dx**  **(N = 58)** | **MCI**  **Dx**  **(N = 134)** | **CL<25:**  **(N = 84)** | **CL>25:**  **(N = 108)** | **APS2 Negative (N = 88)** | **APS2**  **Positive**  **(N=103)** | | **Total**  **(N=192)** |
|  | **APS2** |  |  |  |  |  |  |  |  |
|  | Mean  (sd) | 81.49 (25.24) | 42.14 (32.98) | 21.57 (18.58) | 79.25 (23.20) | 18.85 (12.83) | 83.82 (16.47) | | 53.88 (35.71) |
|  | Min | 7 | 2 | 2 | 11 | 2 | 48 | | 2 |
|  | Max | 100 | 100 | 87 | 100 | 47 | 100 | | 100 |
|  | Missing data | 1 | 0 | 0 | 1 | 0 | 0 | | 1 |
|  | **%p-tau217** |  |  |  |  |  |  |  |  |
|  | Mean  (sd) | 10.84  (5.81) | 5.14  (4.41) | 2.80 (1.69) | 10.02 (5.40) | 2.59 (1.32) | 10.51 (5.14) | | 6.86 (5.52) |
|  | Min | 1.03 | 0.98 | 0.98 | 1.52 | 0.98 | 3.69 | | 0.98 |
|  | Max | 29.91 | 30.68 | 9.35 | 30.68 | 6.59 | 30.68 | | 30.68 |
|  | BLQ | 3 | 40 | 40 | 3 | 43 | 0 | | 43 |
|  | Missing data | 0 | 0 | 0 | 0 | 0 | 0 | | 0 |
|  | **Aβ42/40** |  |  |  |  |  |  |  |  |
|  | Mean  (sd) | 0.0888 (0.0096) | 0.0943 (0.0113) | 0.0983 (0.0107) | 0.0883 (0.0093) | 0.0985 (0.0101) | 0.0877 (0.0094) | | 0.0927 (0.0111) |
|  | Min | 0.0640 | 0.0679 | 0.0724 | 0.0640 | 0.0724 | 0.0640 | | 0.0640 |
|  | Max | 0.1270 | 0.1322 | 0.1322 | 0.1270 | 0.1322 | 0.1270 | | 0.1322 |
|  | Missing data | 1 | 0 | 0 | 1 | 0 | 0 | | 1 |

Mean(sd), minimum and maximum values are reported. APS2 = Amyloid Probability Score 2; BLQ = number of values below the limit of quantitation; CL = centiloid; Dx = Diagnosis; MCI = mild cognitive impairment. Plasma Aβ42/40, %p-tau217 and APS2 values are not reported for one male participant whose plasma was not available for analysis.
